# Convalescent plasma in the management of moderate COVID-19 in India: An open-label parallel-arm phase II multicentre randomized controlled trial (PLACID Trial)

**DOI:** 10.1101/2020.09.03.20187252

**Authors:** Anup Agarwal, Aparna Mukherjee, Gunjan Kumar, Pranab Chatterjee, Tarun Bhatnagar, Pankaj Malhotra, PLACID Trial Collaborators

## Abstract

**Objectives:** Convalescent plasma (CP) as a passive source of neutralizing antibodies and immunomodulators is a century-old therapeutic option used for the management of viral diseases. We investigated its effectiveness for the treatment of COVID-19.

**Design:** Open-label, parallel-arm, phase II, multicentre, randomized controlled trial.

**Setting:** Thirty-nine public and private hospitals across India.

**Participants:** Hospitalized, moderately ill confirmed COVID-19 patients (PaO2/FiO2: 200-300 or respiratory rate > 24/min and SpO2 ≤ 93% on room air).

**Intervention:** Participants were randomized to either control (best standard of care (BSC)) or intervention (CP + BSC) arm. Two doses of 200 mL CP was transfused 24 hours apart in the intervention arm.

**Main Outcome Measure:** Composite of progression to severe disease (PaO2/FiO2< 100) or all-cause mortality at 28 days post-enrolment.

**Results:** Between 22^nd^ April to 14^th^ July 2020, 464 participants were enrolled; 235 and 229 in intervention and control arm, respectively. Composite primary outcome was achieved in 44 (18.7%) participants in the intervention arm and 41 (17.9%) in the control arm [aOR: 1.09; 95% CI: 0.67, 1.77]. Mortality was documented in 34 (14.5%) and 31 (13.5%) participants in intervention and control arm, respectively [aOR) 1.06 95% CI: −0.61 to 1.83].

**Interpretation:** CP was not associated with reduction in mortality or progression to severe COVID-19. This trial has high generalizability and approximates real-life setting of CP therapy in settings with limited laboratory capacity. A priori measurement of neutralizing antibody titres in donors and participants may further clarify the role of CP in management of COVID-19.

**Trial registration:** The trial was registered with Clinical Trial Registry of India (CTRI); CTRI/2020/04/024775.

## Introduction

With few specific therapeutic options available to manage infection with Severe Acute Respiratory Syndrome Coronavirus-2 (SARS-CoV-2), the novel coronavirus disease (COVID-19) presents a unique set of challenges for healthcare providers globally. In addition to non-pharmacological interventions, health systems have responded by devising strategies to manage COVID-19 using repurposed medications and revisiting older strategies, such as using convalescent plasma (CP). CP is a passive immunization strategy which has been used on several occasions in the past century, inspiring clinical expectations that it could emerge as a potential therapy for a disease with no proven, effective interventions.^1^

CP is a source of anti-viral neutralizing antibodies (NAb). Other immune pathways such as antibody-dependent cellular cytotoxicity, complement activation, or phagocytosis are putative mechanisms through which CP may exert its therapeutic effect in COVID-19 patients.^2^ Additionally, anti-inflammatory cytokines, defensins, pentraxins and other immunomodulatory proteins may have a role in alleviating systemic inflammatory response syndrome, the main pathophysiological basis for acute respiratory distress syndrome (ARDS) and mortality from COVID-19 pneumonia.^2^ Historically, CP has been used in viral diseases such as poliomyelitis, measles, mumps and influenza in the pre-vaccine era and more recently, during the Influenza, Ebola, and SARS coronavirus epidemics with varying degrees of success.^3–5^ A recent systematic review on the use of CP in non-COVID-19 severe respiratory viral infections found no mortality benefit.^6^ There is evidence that CP collected from COVID-19 survivors contains Receptor Binding Domain specific antibodies with potent antiviral activity.^7^ However, effective titres of NAb, optimal timing for CP therapy, optimal timing for plasma donation, and the severity class of patients who are likely to benefit from CP therapy remains unclear.

Since the publication of the first case series from China, multiple observational studies have been published, some on pre-print servers, highlighting the usefulness of CP in reducing mortality, hospital stay and viral load in COVID-19 patients.^8–12^ Only two randomized controlled trials on CP use in COVID-19 have been published, one from China and the other from the Netherlands.^13,14^ Both were halted prematurely, the China study due to inadequate patient enrolment and the one from the Netherlands due to a need to redesign the trial based on interim findings. In both studies, no mortality benefit was noted, and the Dutch study raised uncertainties regarding pre-transfusion antibody-status of patients as a potential factor in identifying appropriate candidates for CP therapy.^14^ This uncertainty in the published evidence is reflected in a recent systematic review, which remained undecided on both the safety and effectiveness of CP as a therapeutic option in hospitalized patients of COVID-19.^15^ Meanwhile, CP therapy has received regulatory approval for use in patients in different countries. This has resulted in its widespread adoption in real-world clinical practice, where it is being used to treat COVID-19 patients with a wide spectrum of disease severity.^16,17^ Given these uncertainties, we undertook the current study to determine the effectiveness of using CP in moderately ill COVID-19 patients admitted to hospitals across India in limiting progression to severe disease and determine the associated short-term adverse effects.

## Methods

### Study Design

The PLACID trial was an open-label, parallel-arm, phase II, multicentre, randomized, controlled trial conducted in 39 tertiary care hospitals across. Of these, 29 were teaching public hospitals and 10 were private hospitals spread across 14 States and Union Territories representing 25 cities.

Ethical approval was obtained from the ICMR Central Ethics Committee on Human Research (CECHR-002/2020) as well as from the Institutional Review Boards /Institutional Ethics Committees of all the participating hospitals. The lead author affirms that the manuscript is an honest, accurate, and transparent account of the study being reported; that no important aspects of the study have been omitted; and that any discrepancies from the study as originally planned have been explained. Patient or the public were not involved in the design, conduct, reporting or dissemination plans of our research. The study results will be disseminated to the study via their treating physicians. The study protocol can be accessed at: https://cdsco.gov.in/opencms/opencms/system/modules/CDSCO.WEB/elements/download_file_division.jsp?num_id=NTk2NQ==

### Participants

Patients aged at least 18 years who were confirmed to have COVID-19 based on a positive SARS-CoV-2 RT-PCR test and were admitted to the participating hospitals were screened for eligibility and included if they were moderately ill with either partial pressure of oxygen in arterial blood/fraction of inspired oxygen (PaO2/FiO2) ratio between 200-300 or respiratory rate > 24/min with SpO2 < 93% on room air,^17^ and if matched donor CP was available at the point of enrolment. Pregnant and lactating women, patients with known hypersensitivity to blood products, recipients of immunoglobulin in the last 30 days, patients with conditions precluding infusion of blood products, participants in any other clinical trials and critically ill patients with PaO2/ FiO2 < 200 or shock (requiring vasopressors to maintain a mean arterial pressure (MAP) > = 65 or MAP < 65) were considered ineligible to participate.

Eligible donors were either males or nulliparous females aged between 18-65 years, weighing over 50 kg, who had received a diagnosis of COVID-19 confirmed with a positive RT-PCR test, suffered from symptomatic COVID-19 with at least fever and cough, which had completely resolved for a period of 28 consecutive days prior to donation or a period of 14 days prior to donation with two negative SARS-CoV-2 RT-PCR tests from nasopharyngeal swabs collected 24 hours apart. All routine screening tests including ABO blood grouping, Rh phenotype, complete blood counts, screening for HIV, HBV, HCV, syphilis, malaria and total serum protein were conducted as per the Drugs and Cosmetics (Second Amendment) Rules, 2020.^18^

All participants or their family members or legally authorized representatives were provided with information regarding the trial in a language they were comfortable in, and written informed consent was obtained prior to participant recruitment.

### Randomization and Masking

The randomization sequence was generated using the RALLOC module in STATA v.14 (College Station, Texas, USA) by an independent biostatistician from the Indian Council Medical Research-National Institute of Epidemiology (ICMR-NIE), Chennai, India. A stratified block randomization strategy was used to allocate participants in a 1:1 ratio to receive either CP with the best standard of care (BSC) (intervention arm) or BSC alone (control arm). Stratification was done by sites; block randomization was done with unequal block sizes. After obtaining written, informed consent from an eligible patient, the site investigators screened the participant for recruitment, and called a member of the central trial coordinating team to receive the randomization sequence, ensuring concealment of allocation.

### Procedures

Patients enrolled in the control arm received BSC for COVID-19 in keeping with the institutional protocol, which was dictated by the best available evidence at the time and guidelines for the management of COVID-19 issued by the government health authorities. The range of treatment protocols practised by the participating clinical sites for the management of COVID-19 patients included: antivirals (Hydroxychloroquine, Remdesivir, Lopinavir/Ritonavir, Oseltamivir), broad-spectrum antibiotics, immunomodulators (steroids, Tocilizumab) and supportive management (oxygen via nasal cannula, face mask, non-rebreathing face mask; non-invasive or invasive mechanical ventilation; awake proning). The decision to transfer to the intensive care unit (ICU) was dependent on the policies of the individual trial sites.

The intervention arm received two doses of 200 mL of CP, transfused 24 hours apart, in addition to the BSC. The two plasma units were collected preferably from different donors depending on the availability and ABO compatibility to increase chances of receiving CP with NAb.^19^ If two different donors were not available, both the units were collected from a single donor. CP was collected from recovered COVID-19 patients by centrifugal separation using the apheresis equipment available at the facility after obtaining written informed consent from the donors. At least 20 mL of the donated plasma was stored at −80°C for measurement of SARS-CoV-2 NAb titres, as reliable and approved qualitative and quantitative tests were not available at the time of initiation of the study. Commercial qualitative immunoassays for SARS-CoV-2 antibodies on chemiluminescent immunoassay or enzyme-linked immunosorbent assay platforms approved by the ICMR became available halfway through the trial. Once available, sites were encouraged to use them before collection of CP.

All enrolled participants underwent clinical examination and a range of laboratory investigations including arterial blood gas (ABG) analysis, complete blood count, renal and hepatic function tests, and a coagulation profile on the day of enrolment (Day 0), and subsequently on days 1, 3, 5, 7 and 14. Chest imaging and biomarkers including serum ferritin, Lactate Dehydrogenase (LDH), C-reactive protein (CRP), and D-Dimer were obtained on days 0, 3 and 7. Serum for antibody titre assays was collected on days 0, 3 and 7; these samples were stored −80°C till further analysis at ICMR-National Institute of Virology (NIV), Pune, India. SARS-CoV-2 RT-PCR from nasopharyngeal swabs was repeated on days 3 and 7. All patients were contacted telephonically on day 28 to assess their health status.

Micro-neutralization test for SARS-CoV-2 was performed for determining the NAb titres in stored donor CP and participant serum from day 0, 3 and 7, at the Biosafety Level-3 facility at ICMR-NIV, Pune following standardized methods.^20^ Vero CCL-81-adapted SARS-CoV-2 (strain NIV2020770) isolated at ICMR-NIV, Pune.^21^ The detection range of NAb titers was 1:20 to 1:1280. Values reported as < 1:20 was considered as undetectable NAb titers; values reported as > 1:1280 was considered as 1:1280 for analysis.

### Outcomes

The primary outcome of the study was a composite measure of progress to severe disease (PaO2/FiO2 ratio < 100) any time within 28 days of enrolment or all-cause mortality at 28 days. Primary outcome was considered as “good” if the progress to severe disease or all-cause mortality could be prevented in the 28 days post enrolment and “poor” if not.

The secondary outcomes were clinical improvement and symptom resolution on day 7; variation in fraction of inspired oxygen (Fio2%) on days 1, 3, 5, 7 and 14; total duration of respiratory support during hospitalization and post enrolment duration of respiratory support till day 28 or discharge whichever is earlier; negative conversion of SARS-CoV-2 viral RNA on days 3 and 7; levels of biomarkers on days 3 and 7 post enrolment compared to baseline; requirement of vasopressor support; and clinical improvement on WHO ordinal scale on day 0,1,3,5,7,14 and 28 days.^22^The WHO ordinal scale was not mentioned in the initial study protocol but was added midway through the trial as it was felt that it will be a key endpoint in any future meta-analysis.

Safety outcomes were frequency of minor and serious adverse event (death and invasive mechanical ventilation, hemodynamic instability) within 6 hours of CP transfusion. Relatedness of a serious adverse event with the trial was assessed as per the definitions given by Edwards and Aronson.^23^

### Statistical Analysis

Assuming that 18% of the intervention arm would meet the composite primary outcome under the null hypothesis and 9% under the alternative hypothesis, for power of 80% and significance level of 5%, the sample size was calculated to be 226 in each arm, adding up to 452 participants for the study. The assumption that 18% of the participants in the BSC arm would meet the composite primary outcome was based on the best available evidence at the time of designing the trial.^24^

Descriptive analysis was done by tabulation of data and presentation of continuous variables as mean and standard deviation (SD) or median and interquartile range (IQR), as appropriate, and categorical variables as proportions.

Univariate analysis was done by Chi-squared test to assess the effect of intervention on the composite primary outcome. To adjust for the difference in BSC by trial sites and clustering of patients in some sites, a multilevel logistic regression was done with trial sites taken as the random effect variable along with adjustment for the unequal distribution of diabetes mellitus between the intervention and control arms. Changes in continuous variables like oxygen requirement (FiO2), laboratory parameters (biomarker levels, NAb titres) over the period of hospital stay were compared between both arms by using generalized estimating equations. CRP and D-Dimer levels were categorized as high when levels were > 41.6 mg/L and ≤1 mg/L, respectively.^24,25^ To assess viral clearance, the proportion of participants with SARS-CoV-2 viral RNA negativity was compared between the arms on day 3 and day 7 using the Chi-squared test. The median score on the WHO Ordinal Scale was plotted for the two trial arms for days 0, 1, 3, 5, 7, and 14.

Post-hoc subgroup analysis was done to compare the composite outcome between participants who received CP with detectable NAb and control arm. Patients receiving CP with NAb titre > 1:80 were compared with control for the primary outcome. Stratified analysis was done for the primary outcome between intervention and control arms based on strata such as detectable NAb at enrolment in participants, and duration of symptoms at enrolment. To assess the effect of transfusing CP early in the disease, subgroup analysis for the composite outcome was conducted in participants who had symptoms for less than or equal to 3 days at enrolment.

An intention-to-treat analysis (ITT) was done after imputing the missing composite outcomes as progression to severe disease or mortality. Data were collected in structured paper case record forms (CRF) and then entered in the Research Electronic Data Capture system (REDCap, Version 8.5 Vanderbilt University, Nashville, TN). Data analysis was done using STATA v14 (College Station, Texas, USA). An independent Data and Safety Monitoring Board (DSMB) oversaw the study. The trial protocol was registered with the Clinical Trial Registry of India (CTRI); registration number CTRI/2020/04/024775.

## Results

A total of 1210 patients admitted across 39 trial sites were screened between 22^nd^ April to 14^th^ July 2020; 464 patients were randomized into intervention or CP +BSC arm (n = 235) and control or BSC arm (n = 229). Outcome at 28 days was not available for 2 patients (one each in intervention and control arm) who were lost to follow-up after discharge from hospital; 9 patients (5 in intervention and 4 in control arm) withdrew consent after randomization. All 464 patients have been included in the intention-to-treat analysis.

Baseline demographic and clinical characteristics were similar across the trial arms, except for prevalence of diabetes mellitus and cough. (Tables 1 and 2). Patient management across the trial arms was similar except for CP therapy. (Table 2)

**Table 1:** Demographic and baseline characteristics of the study participants, n = 464

|  | **Best Standard of Care arm, n=229** | **Convalescent Plasma arm, n=235** |
| --- | --- | --- |
| **Age in yrs, Median (IQR)** | 52 (41,60) | 52 (42, 60) |
| **Gender, male, n(%)** | 177 (77.3%) | 177 (75.3%) |
| **BMI, kg/m<sup>2</sup>, mean (SD)** | 26.1 (4.2%) | 26.2 (4.3%) |
| **Comorbidities, n(%)** | 147 (64.2%) | 167 (71.1%) |
| **Diabetes mellitus** | 87 (38.2%) | 113 (48.1) |
| **Hypertension** | 81 (35.5%) | 92 (39.1%) |
| **Coronary artery disease** | 17 (7.4%) | 15 (6.4%) |
| **Obesity** | 17 (7.4%) | 16 (6.8%) |
| **#Tuberculosis** | 10 (4.4%) | 9 (3.8%) |
| **Chronic kidney disease** | 9 (3.9%) | 8 (3.4%) |
| **Chronic obstructive pulmonary disease** | 7 (3.1%) | 8 (3.4%) |
| **Cerebrovascular disease** | 1 (0.4%) | 3 (1.3%) |
| **Liver Cirrhosis** | 2 (0.9%) | 0 |
| **Cancer (History)** | 0 | 1 (0.4%) |
| **Ever smoker, n(%)** | 18 (7.9) | 19 (8.1) |
| **Blood Group, n(%)** |  |  |
| **A** | 51(22.3%) | 55(23.4%) |
| **B** | 83(36.3%) | 87(37.1%) |
| **O** | 83(36.2%) | 79(33.6%) |
| **AB** | 12(5.2%) | 14(5.9%) |
| **Symptom onset to admission, days, Median (IQR)** | 4 (3,7) | 4 (3,7) |
| **Symptom onset to enrolment, days, Median (IQR)** | 8 (6,11) | 8 (6,11) |
| **\*Detectable Neutralisation antibody titre, n(%)**<br>**n=418** | 163 (80.3) | 185 (86.1) |

\*Data not available for all randomized patients # Only 2 patients had active TB, one in each arm
BMI: body mass index; IQR: interquartile range

**Table 2:** Clinical and laboratory parameters of the trial participants at baseline and medications received during hospitalization, n = 464

|  | **Best Standard of Care arm (n=229)** | **Convalescent Plasma arm (n=235)** |
| --- | --- | --- |
| **Shortness of Breath, n(%)** | 208 (90.8%) | 215 (91.5%) |
| **Fever, n(%)** | 85(37.1%) | 77(32.8%) |
| **Cough, n(%)** | 167(72.9%) | 149(63.4%) |
| **Fatigue, n(%)** | 182/229 (79.5%) | 183/234 (78.2%) |
| **X-ray findings, n(%), n=432** |  |  |
| Ground glass opacity | 29 (13.5%) | 27 (12.4%) |
| Local patchy shadows | 9(4.2%) | 12(5.5%) |
| B/L patchy shadows | 139(64.9%) | 140(64.2%) |
| Interstitial abnormalities | 4(1.9%) | 3(1.4%) |
| B/L white out lung | 2(0.9%) | 2(0.9%) |
| **SpO2 (%) on room air, mean (SD), n=420** | 88.5(4.1) | 88.1 (4.4) |
| **FiO2 required to maintain SpO2 >92%, mean (SD), n=462** | 37.4 (11.4) | 39.04 (13.1) |
| **PaO2/FiO2, mean (SD)** | 251.6 (39.5) | 255.4 (42.1) |
| **Haemoglobin (g/dl), mean (SD)** | 12.5 (1.8) | 12.5 (2.1) |
| **WBC count (Cells /mm<sup>3</sup>), median (IQR)** | 8500 (6500,11200) | 8480 (6110,11460) |
| **Neutrophil-Lymphocyte Ratio (NLR), median (IQR)** | 5.5 (3.4, 9.4) | 5.5 (3.5, 10) |
| **Ferritin, ng/mL, Median (IQR), n=448** | 539.5 (328.3, 873) | 529.8 (278.6, 956) |
| **LDH, IU/L, Median (IQR), n=450** | 458.6 (342.5, 638.5) | 473.5 (335, 661) |
| **High CRP, n(%), n=395** | 97 (50%) | 100 (49.7%) |
| **High D-Dimer, n(%), n=399** | 68 (34.7%) | 82 (40.4%) |
| **WHO Ordinal scale, n(%), n=463** |  |  |
| **4** | 181(79.0%) | 180 (76.9%) |
| **5** | 47(20.5%) | 54(23.1%) |
| **6** | 1(0.4%) | 0 |
| **Medications, n(%)** |  |  |
| **Hydroxychloroquine** | 155 (68.3%) | 159 (67.7%) |
| **Remdesivir** | 13 (5.9%) | 7 (3.2%) |
| **Lopinavir/Ritonavir** | 30 (13.6%) | 36 (16.1%) |
| **Methylprednisolone** | 114 (49.8%) | 123 (52.3%) |
| **Dexamethasone** | 30 (13.1%) | 23 (9.8%) |
| **Hydrocortisone** | 5 (2.2%) | 4 (1.7%) |
| **Tocilizumab** | 26 (12.7%) | 16 (7.2%) |
| **Heparin (UFH/LMWH)** | 170 (76.6%) | 178 (77.7%) |
| **Azithromycin** | 140 (65.1%) | 156 (69.3%) |
| **Intravenous Immunoglobulin** | 0 | 1 (0.4%) |
| **Other Antibiotics** | 196 (87.5%) | 204 (87.5%) |

High CRP: Levels above 41mg/L; High D-dimer: Levels above 1mg/L
FiO2: fraction of inspired oxygen; PaO2: Partial pressure of Oxygen in arterial blood; WBC: White Blood Cells; LDH: lactate dehydrogenase; CRP: C-reactive protein; WHO Ordinal scale: World Health Organization ordinal scale; UFH: unfractionated heparin; LMWH: Low molecular weight heparin.

CP was collected from 433 donors; CP of 262 donors was finally used for the trial. Most of the donors were male (94.3%), with mean (SD) age of 34.3 (9.3) years. The median (IQR) disease duration was 6 (3,11) days and most of the donors (94.2%) had mild disease. Nearly two-thirds (63.6%) of the donors had a NAb titre of more than 1:20 with a median (IQR) titre of 1:40 (1:30,1:80). Plasma was donated after a median (IQR) of 41 (31,51) days from COVID-19 diagnosis by RT-PCR.

There was no difference in the primary outcome across the trial arms on ITT analysis. Thirty-four patients (14.5%) died in the intervention arm and 31 (13.5%) in the control arm; adjusted odds ratio (aOR) was 1.06 [95% CI: 0.61 to 1.83]. Seventeen patients each progressed to severe disease in both arms [7.2% vs. 7.4%; aOR: 1.04; 95% CI: 0.51 to 2.11]. The composite outcome was achieved in 44 (18.7%) patients in the intervention arm and 41 (17.9%) in the control arm [aOR: 1.09; 95% CI: 0.67 to 1.77]. (Table 3)

**Table 3:** Comparison of primary outcomes between intervention and control arms

| | Best Standard of Care arm<br>N=229 | Convalescent Plasma arm*<br>N=235 | Convalescent Plasma arm (with detectable NAbs in CP)*<br>N=160 | Convalescent Plasma arm (with NAbs≥1:80 in CP)*<br>N=67 | Convalescent Plasma arm (with undetectable NAb in CP)*<br>N= 64 |
| --- | --- | --- | --- | --- | --- |
| All-cause mortality at 28 days | 31(13.5%) | 34 (14.5%) | 22 (13.8%) | 10 (14.9%) | 12 (18.7%) |
| aOR (95%CI) |  | 1.05 (0.61,1.83) | 0.93 (0.49, 1.70) | 0.93 (0.39, 2.21) | 1.49 (0.68, 3.23) |
| Progression to severe disease (P/F ratio <100) | 17 (7.4%) | 17 (7.2%) | 11 (6.8%) | 6 (8.9%) | 6 (9.4%) |
| aOR (95%CI) |  | 1.04 (0.51, 2.11) | 0.91(0.41, 2.02) | 1.25 (0.45, 3.41) | 1.33 (0.49, 3.59) |
| Composite outcome | 41 (17.9%) | 44 (18.7%) | 27 (16.9%) | 12 (17.9%) | 13 (20.3%) |
| aOR (95%CI) |  | 1.09 (0.67, 1.77) | 1.04 (0.61,1.78) | 0.92 (0.43,1.97) | 1.19 (0.59, 2.42) |

aOR: adjusted odds ratio; NAb: neutralizing antibodies; CP: Convalescent plasma
\*All comparisons were done with best standard of care, adjustment done for presence of diabetes mellitus and trial site

The proportion of patients with resolution of shortness of breath and fatigue at day 7 were higher in the intervention arm, whereas fever and cough resolution were not different in the two groups. (Table 4) The median (IQR) absolute reduction in Fio2% on days 3 [5 (0,12) vs. 3.7 (0,9), p = 0.04], and 5 [9 (3,17) vs. 7 (0,14), p = 0.04] from the day of enrolment was significantly more in the intervention as compared to control arm. At 7 [CP+BSC: 11 (6,18) vs. BSC: 9.5 (6,18), p = 0.48;] and 14 days [CP+BSC: 14 (9,19) vs. BSC: 12 (6,19), p = 0.18;] post enrolment, the change in FiO2% from day of enrolment was similar between the two arms. The negative conversion of SARS-CoV-2 viral RNA at day 7 post-enrollment was significantly higher in the CP arm compared to the BSC arm. (Table 4)

**Table 4:** Comparison of secondary outcomes between intervention and control arms

|  | Basic Standard of Care arm, N=229 | Convalescent Plasma arm, N=235 | P Value |
| --- | --- | --- | --- |
| **Resolution of** shortness of breath on day7, n (%) | 119 (65.7%) | 140 (76.5%) | 0.02 |
| **Resolution of** fever on day7, n (%) | 65 (91.5%) | 67 (98.5%) | 0.06 |
| **Resolution of** cough on day7, n (%) | 111 (75.5%) | 102 (79.7%) | 0.4 |
| **Resolution of** fatigue on day7, n (%) | 92 (60.1%) | 114 (73.1%) | 0.02 |
| **Absolute change in Fio2 on day 3** | 3.7 (0,9) | 5 (0,12) | 0.04 |
| **Absolute change in Fio2 on day 5** | 7 (0,14), | 9 (3,17) | 0.04 |
| **Absolute change in Fio2 on day 7** | 9.5 (6,18), | 11 (6,18) | 0.48 |
| **Negative conversion of SARS-CoV-2 viral RNA on day 3, n=350, n(%)** | 67 (36.2%) | 82 (43.4%) | 0.15 |
| **Negative conversion of SARS-CoV-2 viral RNA on day 7, n=374, n(%)** | 94 (54.6%) | 121 (67.9%) | 0.01 |
| **Total days of hospital stay, Median (IQR)** | 13 (10,18) | 14 (10,19) | 0.51 |
| **Total days of respiratory support, Median, IQR** | 10 (6,13) | 9 (6,13) | 0.67 |
| **Days of respiratory support post-enrolment, Median (QR)** | 6 (4,10) | 6 (3,9) | 0.46 |
| **Invasive mechanical ventilation during hospital stay, n(%)** | 20 (8.9%) | 18 (7.8%) | 0.68 |
| **Vasopressor support after enrolment, n(%)** | 8 (3.6%) | 10 (4.4%) | 0.7 |

All changes are measured from day of enrolment; FiO2: Fraction of inspired oxygen

Total and post-enrollment duration of respiratory support, proportion of patients requiring invasive ventilation, and proportion of patients requiring vasopressor support were not different between the trial participants in two arms (Table 4). Among the 38 participants who required invasive ventilation, only two participants survived till 28 days post enrolment. The average levels of inflammatory markers over a period of 7 days from enrolment, showed no statistically significant difference between the two arms [LDH (β = 8.9, 95%CI-0.53, 70.9), ferritin (β = 30.59, 95%CI −51.36, 112.55). There was no difference in WHO ordinal scale for clinical improvement between the two arms at any time point of observation.

In the trial participants who received CP, minor adverse events of pain in local infusion site, chills, nausea, bradycardia and dizziness was reported in one patient each. Fever and tachycardia were reported in three patients each. Dyspnoea and intravenous catheter blockage were noted in two participants each. Mortality was assessed as possibly related to CP transfusion in 3 patients (1.3%).

Stratified analysis was done by symptom duration at enrolment. The composite outcome was not different between the intervention and control arms for patients with symptom duration less than or equal to 3 days at enrolment [n = 24, aOR: 0.59 (95% CI: 0.28, 1.24)]. Titres of NAb in transfused CP were available in 224 out of 235 participants in the intervention arm; 160 (71.4%) participants received at least one unit of CP with detectable NAb. The primary outcome was not different between the subgroups of intervention arm patients receiving CP with detectable NAb titres (n = 160) or CP with NAb titre ≥1:80 (n = 68) or CP with no detectable NAbs (n = 64) and the control arm (Table 3).

NAb titres were measured in 418 trial participants; 348 (83.2%) had detectable NAb at enrolment. The median (IQR) NAb titre at enrolment was 1:90 (1:30,1:240). In enrolled participants with detectable NAb at baseline, the CP and control groups were not different in terms of the composite primary outcome [29 (15.7%) vs. 27 (16.6%), aOR: 1.01 (95%CI: 0.56,1.81)]. In patients with undetectable NAb at baseline (n = 70), no difference of composite primary outcome could be discerned [9 (30%) vs. 10(25%), aOR: 1.75 (95%CI: 0.41,7.51)].

## Discussion

The PLACID trial results indicate that there was no difference in 28-day mortality or progression to severe disease among moderately ill COVID-19 patients treated with CP along with BSC compared to BSC alone. Additionally, there were no differences in outcomes between study participants receiving CP with detectable NAb titres compared to BSC alone; or between those receiving CP with NAb titres more than or equal to 1:80 and those receiving BSC alone. CP therapy was associated with a higher resolution of symptoms like shortness of breath and fatigue on day 7. CP use was also associated with reduction in FiO2 requirement on day 3 and 5 but not in the duration of respiratory support. There was a higher proportion of negative conversion of viral RNA on day 7 post enrolment in intervention arm. However, it did not demonstrate anti-inflammatory properties as we could not detect any difference in the levels of inflammatory markers such as ferritin, CRP, D-Dimer or LDH between the two arms.

A recent Cochrane review, including 20 studies [1 RCT, 3 controlled non-randomized study of intervention (NRSI), 16 non-controlled NRSIs], concluded that there is uncertainty regarding effectiveness of CP in improving mortality or clinical improvement in COVID-19 patients.^15^ In a randomized controlled trial conducted from China, Li and colleagues studied 103 patients with severe and life-threatening COVID-19 and reported no effect on time to clinical improvement. Their subgroup of 45 patients with severe disease, similar to patients with moderate disease in our study, demonstrated increased clinical improvement in their CP group.^13^ The ConCOVID trial from the Netherlands, prematurely terminated after enrolling 86 patients, could not find any effect on mortality at 60 days, hospital stay or 15-day disease severity.^14^ A large observational study advocated the usefulness of CP in COVID-19, stating that the 7-day and 30-day mortality were lower in those who received CP within 3 days of symptom onset. However, the absence of a controlled comparison weakens these findings as evidence of efficacy.^26^ Although underpowered, we did not find any benefit of administration of plasma within 3 days of symptom onset.

Our results mirror the experience of the ConCOVID trial, where 79% of the participants had detectable antibodies at baseline.^14^ On the other hand, the NAb titres in CP in our study was similar to the results by Long et al, who found 13-40% patients turned seronegative in the early convalescent phase.^27^ In a 175-patient series, Wu et al documented that 30% of patients generated very low levels of NAb with titres correlating to increasing age and disease severity.^20^ We found participants had higher antibody positivity and median NAb titres than CP donors. The difference in age and severity of illness, with donors being younger and having milder disease, could have driven this difference. While all COVID-19 survivors were encouraged to donate plasma, an overwhelming majority of the donors were only mildly sick, young survivors. Recovered patients who had moderate or severe disease were generally reluctant to return to hospitals for plasma donation. This has major implications for obvious operational reasons as CP therapy for COVID-19 gets scaled up as a component of COVID-19 management strategy not only in India but also globally.

There was no difference in NAb titres in the two arms despite CP transfusion. This suggests that there may not be any benefit of CP collected from young mild COVID-19 recovered donors to moderate to severely sick elderly patients who have a robust antibody response.

The early conversion of viral RNA negativity in intervention arm aligns with published evidence, and further supports the fundamental hypothesis that CP exerts virus neutralization effects.^11,13^ However, the goal of getting better clinical outcomes remained elusive for us. We did not find evidence to support the immunomodulator functions of CP as we could not demonstrate differences in the levels of inflammatory markers. This could potentially explain why CP had no difference in primary outcome despite early SARS-CoV-2 viral RNA negativity.

CP transfusion was deemed to be safe in our study; minimal non-life-threatening adverse events were reported. Three deaths could possibly be related to CP transfusion, which is comparable to the other larger reports of safety of CP use in COVID-19.^12^ Possible is defined here as a clinical event which occurred within 6 hours of CP transfusion, but could also be explained by worsening COVID-19.^23^

The PLACID trial was conducted to generate context-specific evidence relevant to all stakeholders, including policymakers, healthcare providers, and patients. To achieve this, expression of interest was invited from hospitals across the country and not limited to a few centres of excellence. Hospitals were chosen both from the public and private sectors, lending heterogeneity in infrastructure, and wide socio-cultural-economic representation of study participants, with a large range of co-morbidities and presenting features. The BSC represented the care standards likely to be provided in a real-world setting. While this approach could have impacted the internal comparability across sites, we feel this lends the trial more generalizability, approximates real-world scenarios more closely and places our study closer to the pragmatic trials across the methodological spectrum of clinical trials.^28^

The PLACID trial has several limitations. Being an open-labelled study, it was susceptible to anchoring bias in treating physicians in outcome ascertainment. This may be reflected in the higher resolution of subjective symptoms like shortness of breath and fatigue noted in intervention arm. The trial was conducted in 39 hospitals across the country with some level of heterogeneity across the trial sites in terms of BSC and participant enrolment. The biomarker assays for ferritin, LDH, CRP and D-dimer were conducted with products from different manufacturers. Also, as the pandemic was in different stages across the country, the enrolment strength varied from site to site with a possibility of selection bias arising due to clustering of enrolment. We attempted to alleviate this bias by statistically adjusting the primary outcome for trial sites. Further, the discharge criteria for COVID-19 was based on government guidelines and not on clinician discretion and patient condition, hence we chose not to analyse this as a secondary outcome. We could not measure the antibody titres in CP before transfusion, since validated, reliable, commercial tests for qualitative or quantitative antibody measurement were not available at the time of commencement of this trial. However, this remains a close approximation of the manner in which CP therapy has largely been deployed in regions with limited laboratory capacity.

Use of CP as a treatment modality for COVID-19 has received authorisation for off-label use in India.^29^ This authorization has been paralleled by questionable practices such as calls for donors on social media, and the sale of CP on the black market with exorbitant price tags in India.^30^ Additionally, although CP is a safe therapeutic modality, plasmapheresis, plasma storage and NAb measurement are all resource-intensive processes, with limited number of institutes in the country having the capacity to undertake these activities in a quality-assured manner.

Although the use of CP seemed to improve resolution of shortness of breath and fatigue, reduce FiO2 requirement in the first week and led to higher negative conversion of viral RNA, this did not translate into reduction in 28-day mortality or progression to severe disease in moderate COVID-19 patients. Areas of future research could include effectiveness of CP among NAb naïve patients and the use of CP with high NAb titres.

## Contributors

### Patient enrollment, conduct of study, clinical care and data collection

Madras Medical College, Chennai: BL, SS, SAMK, VR, AS, PB, RSUM, RJ, SR.

SMS Medical College, Jaipur: SB, SB, AS, AP, AH, GR.

Sir H.N Reliance Foundation Hospital and Research Centre: VK, KK, JR, DR, EP, NB, MHP, RJV.

Sri Aurobindo Institute of Medical Science, Indore: RD, SP, AT, SJ, RK.

Smt. NHL Municipal Medical College, Ahmedabad: JRK, NNS, NMS, HMP, CKS, MNP, SS, SHS, TM, VRB. Topiwala National Medical College and BYL Nair Hospital: RDS, KJ, FE, SA, RB, AMN, TM, VK, RW, NV. Gandhi Medical College, Secunderabad: RRM, BTC, AVS, AKM, KH, KN, KS, TRC, KTR, JV.

Government Institute of Medical Science, Noida: SS, RU, SB, RP, SS, BRG.

Gandhi Medical College, Bhopal: SD, RS, PD, RM, DC, JL, UMS, JLM.

ABVIMS & RML Hospital, New Delhi: KC, AS, VK, RK, PK, BPA, KKG, AG, PS, SD.

Satguru Pratap Singh hospitals, Punjab: AJ, MJJ, ASD, RK, NS, NK, DK, RK, RM, GS, JK, RPS, RB

Kasturba Hospital: SP, OS, JS, MD, SU.

Rajarshree Chhatrapati Shahu Maharaj Govt Medical College: VAF, VB, RM, SY

All Institute of Medical Sciences, Jodhpur: SM, AB, MKG, GKB, VN, PBA, MN, PS, RN

Post Graduate Institute of Medical Education and Research, Chandigarh: NSK, PM, RS, MPS, NS, SS, RH, VS, LNY, PVML

All Institute of Medical Sciences, Patna: NS, DB, NK

Byramjee Jeejeebhoy Medical College, Pune: MT, SS, NK, SS, LN, SJ, RK, SG

ESIC Medical College and Hospital, Faridabad: NS, NV, AD; Clinical Development Services Agency: MB, NW Smt. Kashi Bai Navale Medical College: SB, SD, VW, AK, TY

Karnataka Institute of Medical Sciences, Hubballi: RSK, PR, KY, PG, VM, MS, MHN

Lady Hardinge Medical College and SSK Hospital, New Delhi: AG, RS, SP, AP, PG, SS

King George’s Medical University, Lucknow: DHR, CT, SP, PM, AW, VK

Byramjee Jeejeebhoy Medical College, Ahmedabad: KU, NB, NS, MS, TP

Mahatma Gandhi University of Medical Sciences Technology, Jaipur: RMJ, AJ, SS, PR, NG

Government Medical College, Surat: TCP, MGS, JP, YRS, MJ

GMERS Medical College and Hospital, Gotri, Vadodara: VG, MS, RR, IN

Sumandeep Vidyapeeth and Dhiraj Hospital, Vadodara: PRJ, ADS, GY, AJ, RKG

Sri Venkateshwara Institute of Medical Sciences, Tirupati: KVSB, BSB, AM, BV, KCS

Kurnool Medical College, Kurnool: SD, KN, CA, GB, RRK, PC

Madurai Medical College, Madurai: MN, MS, DPK, FR

Government Medical College, Bhavnagar: SJP, PJA, KPJ, PHS, MB

MGM Indore: MB, AY, MG, NR, DC

Poona Hospital and Research Centre: VKK, DP, SM, CDS, VT

Super Specialty Pediatric Hospital and Post Graduate Teaching Institute, Noida: SA, DKG, SD

SN Medical College, Agra: NC, ASC

Christian Medical College, Vellore: JJM, SK, DD

Aditya Birla Memorial Hospital: RS, VD, YA, SA

RD Gardi Medical College, Ujjain: AP, MP, AS

Seth GS Medical college and KEM Hospital, Mumbai: JS, KJ, SB; National Institute of Immunohaematology: MM, RMY

Lokmanya Tilak Municipal Medical College & General Hospital, Mumbai: NDK, YAG, LN, SM

### Study design, data analysis, data interpretation and manuscript writing team

AA, AM, GK, PC, TB, SD, PM.

### Data Management Team

KK, RS.

### Generation of randomization sequence

VSK.

### Central Implementation Team

AA, AM, GK, AT.

### Laboratory Analysis Team

GD, SS, RG, AS, DP, CP, SS, KJ, HK, PDY, GS, PA.

## Data Availability

Individual patient level data, collected in connection with the PLACID Trial, along with a data dictionary defining the variable fields has been developed and is available with the corresponding author. De-identified participant level data with data dictionary, or a sub-set thereof, may be made available upon written request to the corresponding author. Additional documents, including the study protocol, statistical analysis plan have been made available as supplementary files within the original submission. Results of secondary or sub-group analyses will be provided on request. Data will be made available, upon request once the trial is published. Data requests should be accompanied by a brief proposal outlining the analysis plan, which may be carried out with investigator support. A signed data access agreement may be needed to ensure data safety and compliance with national rules regarding data sharing.

## Acknowledgement

We wish to acknowledge the following people:

Dr. Santanu Kumar Tripathi, Colonel Rajat Jagani, Prof. L Jeyaseelan, Dr. MS Jawahar and Dr. Suman Kumar Pramanik were members of the independent Data and Safety Monitoring Board.

Dr. S. Devika for helping with study images.

Mr. Jagdish Rajesh, Dr. R Lakshminarayanan and Ms Cosmic Priyanka Singh for their constant administrative support at ICMR Headquarters.

Ms Rekha Dubey, Dr. Rajshekhar Iyer, Dr. Naveen Dutt, Dr. Maya Gopalakrishnan, Dr. Deepak Kumar, Dr. PK Singh, Dr. CM Singh, Dr. Urmika Dholiya, Dr. Vrushti Doshiyad, Dr. Mudita Ravani, Dr. Anand Zachariah, Dr. Binila Chacico, Dr. Dheeraj Kumar, Dr. Shalini Shukla, Dr. Indal Chauhan, Dr. Aruna Kumar, Dr. A.K Shrivastava, Dr. MD Dawood Suleman, Dr. K. Padmamalini, Dr. B. Sheshadri, Dr. Iqbal Ahmed, Dr. Vijay Shah, Dr. Kedar, Dr. Tejas Patel, Dr. Tarang Gianchandani, Dr. Chetan B Bhatt, Ms. Ayushi Gianchandani, Dr. Hemant Deshmukh, Dr. Swait Kulkarni, Dr. Gita Nataraj, Dr. Virendra Atam, Professor Amita Jain, Professor Mahendra Lal Brahmbhatt, Dr. Ramalingappa Antaratani, Dr. Iswar S.Hasabi, Dr. Sachin Hosakatti, Dr. Vikas Chandra Swarnkar, Dr. Ganesh Narain Saxena, Dr. Ashina Singla, Dr. R. Chitraa, Dr. CS Sripriya, Dr. J Bharathi Vidhya Jayanthi, Dr. K Ramadevi, Dr. M Chitra, Dr. G Shanthi, Dr. Jagat Ram, Dr. Govardhan Dutt Puri, Ms Richa Pandey, Dr. KK Talwar, Dr. Bishav Mohan, Dr. Sandeep Kaushal, Dr. Loveena Oberoi, Dr. Lalit Kumar Garg, Dr. Rupinder Bakshi, Dr. Kanwaljit Kaur, Dr. Sarabjeet Sharma, Dr. Harjot Kaur, Dr. Gurinder Mohan, Dr. Sunil Chawla, Dr Sarajeet Sharma, Professor Neelam Marwaha, Dr. Ravinder Garg, Dr. Shipa Arora, Dr. Avneet Kaur, Dr. Rajesh Bhaskar, Dr. Gagandeep Singh Grover, Dr. Shilpa Arora, Dr. Sandeep Kaushal, Dr. Rama Gupta, Dr. Prabhat Mehta, Dr. Sushmita Tripathi, Dr. Sachin Dhanrale, Dr. Shreetoma Datta, Dr. Snehal Bachuwar, Dr. Radheshyam Chejara, Dr. Uday Singh Meena, Dr. Girraj Prasad Mathuria, Dr. Dharmendra Kumar Singh, Dr. Prabhat Aggarwal, Dr. Nita Radhakrisnan, Dr. Ankur Goyal, Dr. J.Sangumani, Dr.Senthil and Dr. M.Suresh Kumar helped us enrol participants at the study sites.

Dr. Nitin M Nagarkar, Dr. MD Sabah Siddiqui, Dr. Ramesh Chandrakar, Dr. CH Srinivasa Rao, Dr. Rajiv Kumar B, Dr. Emine A Rahman, Dr. Abhishek B, Dr. Anusha Cherian, Dr. Tamilarasu Kadhiravan, Dr. Maanas Bhaskar, Dr. Aseem K Tiwari, Dr. Geet Aggarwal, Dr. Swati Pabbi, Dr. Arti Trivedi, Dr. Krupal Pujara, Dr. Shailesh Mundhava, Dr. Sudha Ramalingam, Dr. A Murali, Dr. Karthikeyan, Dr. Anjali Sharma, Dr Nandini Duggal, Dr. Mala Chabbra, Dr. Taruna Bansal, Dr. Anupam Verma, Dr. Rahul Katharia, Dr. R.K Dhiman, Dr. S. Alagesan, Dr. SA Manimala, Dr. J Ravishankar, Dr. Inbanathan.J, Dr.Vikas Laxman, Dr. Bharath.M.S, and Dr.Madhukumar.R for their constant support to the study.

## Copyright for publication

The Corresponding Author grants on behalf of all authors a worldwide licence to the Publishers and its licensees in perpetuity, in all forms, formats and media (whether known now or created in the future), to i) publish, reproduce, distribute, display and store the Contribution, ii) translate the Contribution into other languages, create adaptations, reprints, include within collections and create summaries, extracts and/or, abstracts of the Contribution, iii) create any other derivative work(s) based on the Contribution, iv) to exploit all subsidiary rights in the Contribution, v) the inclusion of electronic links from the Contribution to third party material where-ever it may be located; and, vi) licence any third party to do any or all of the above.

## Data Sharing Statement

#### What is already known on this topic

Till date, multiple small case-series, one large observational study with over 35,000 patients, and two randomized clinical trials have been published on the utility of convalescent plasma in management of COVID-19. Whilst the observational studies suggested clinical benefits in recipients of convalescent plasma, the trials were stopped early, and they failed to ascertain any mortality benefit associated with convalescent plasma therapy in COVID-19 patients.

#### What this study adds

In settings with limited laboratory capacity, convalescent plasma does not reduce 28-day mortality or progression to severe disease in moderately ill, hospitalized COVID-19 patients. It was associated with resolution of shortness of breath fatigue on day 7, reduction in supplemental oxygen requirement on day 3 and 5 and early negative conversion of SARS-CoV-2 viral RNA. The effectiveness of convalescent plasma as a potential therapeutic modality for moderately ill COVID-19 patients is limited.

